# MuPET (Multiphysics PET): Bridging Computational Mass Transport Physics and Medical Imaging for Synthetic Positron Emission Tomography Data Generation

**DOI:** 10.1101/2025.07.31.25332512

**Authors:** Sai Kiran Kumar Nalla, Quentin Maronnier, John A. Kennedy, Sebastian Uppapalli, Olivier Caselles

## Abstract

**Purpose:** Robust positron emission tomography (PET) research increasingly requires reliable ground-truth activity distributions, yet clinical data and physical phantoms cannot provide them fully. This work introduces MuPET (multiphysics PET), a computational framework that bridges mass-transport physics simulation with image domain noise modeling to generate realistic, synthetic PET images.

**Methods:** Tracer transport and radioactive decay within a custom phantom geometry were modeled via decay diffusion physics. The simulated activity concentrations were converted into PET equivalent count maps, followed by a pseudo-Poisson noise model calibrated to scanner-specific characteristics. MuPET generated images were compared with experimental PET acquisitions and a conventional synthetic lesion insertion pipeline. VOI (volume of interest) and ROI (region interest) data extraction was performed on all the data. The quantitative metrics included the recovery coefficient (RC), target contrast ratio (TCR), coefficient of variability (COV), and activity profile.

**Results:** The simulated effective activities matched the decay-corrected measurements within 0.4–1.2%, confirming high physics-model fidelity. The optimized noise model achieved COV values (3.9–4.0%) comparable to those of the acquisitions (3.8–3.9%). MuPET demonstrated strong agreement with the experimental profiles and quantitative metrics, with mean biases within 10% of the acquisition.

**Conclusion:** MuPET establishes a fast (approximately 15 min), physically grounded pipeline that combines transport modeling with PET image simulation, enabling generation of realistic datasets. This approach provides a practical foundation for future dynamic PET simulations.

*Graphical Abstract:* 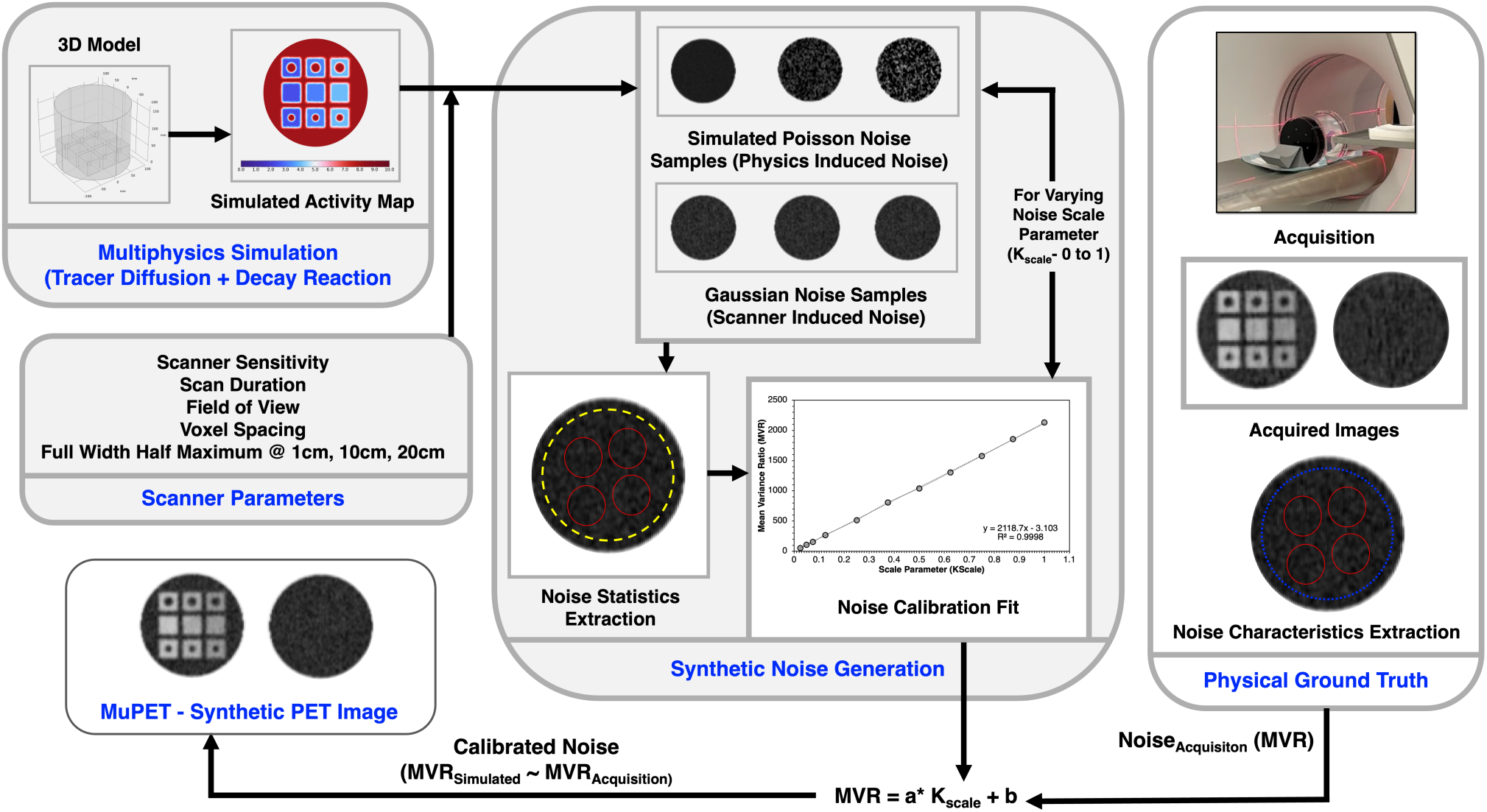

## Introduction

Positron emission tomography (PET) in combination with computed tomography (CT) imaging offers unique insights for the early diagnosis of cancer **[1–3]**. Robust quantitative PET analysis, radiomics pipelines, and data-driven imaging methods increasingly require reliable ground-truth activity distributions to benchmark performance **[4–6]**. However, clinical PET acquisitions alone cannot provide true ground truth data due to biological variability and ethical limitations. Traditionally, simpler systems such as standardized phantoms are designed for benchmarking quantitation and image quality tests across generations of scanners to generate ground truths **[7]**. However, current phantoms lack the customizability necessary to mimic anatomically realistic conditions accurately **[8–10]**.

To address this need, PET simulation frameworks have been developed, ranging from analytical models to highly detailed Monte Carlo tools such as SimSET or GATE **[11,12]**. These simulators replicate detector physics and photon transport with high fidelity. However, their computational expense and complexity make them impractical for rapid iteration and large-scale dataset generation. Projection-domain lesion-insertion methods such as DUETTO **[13,14]**, PETSTEP (PET simulator of tracers via emission projection) **[15]**, SMART PET (SiMulAtion and ReconsTruction) **[16]** and FAST PET (fast analytical simulator of tracers) **[17]** offer faster alternatives. They inserted synthetic activity directly into sinograms prior to vendor reconstruction. While these approaches can approximate clinical acquisition pipelines, they rely on a baseline PET scan or ideal activity source map with additional noise modeling. Models often combine Poisson, pseudo-Poisson, or gamma-based assumptions for replicating stochastic noise **[18–22]**. However, these approaches remain constrained by the availability of baseline PET scans and offer limited flexibility for creating synthetic data in new geometries or dynamic conditions.

In parallel, three-dimensional (3D) printing has emerged as a key experimental tool for creating more realistic customizable anthropomorphic phantoms at a lower cost **[23–27]**. This facilitates the rapid prototyping of complex models, allowing for the inclusion of tissue heterogeneity and detailed lesion morphologies. The adaptability of 3D-printed phantoms across facilities for standardization and harmonization studies has also been established **[10,28,29]**. However, this alone cannot meet the demands of scalable ground-truth generation, since every new design iteration must still be physically manufactured, filled, and imaged. Thus, the limitation underscores the broader need for computational models to create synthetic PET data for ground truths. With the advent of digital twins various medical domains have long relied on physics-based modeling, including cardiovascular modeling **[30,31]**, perfusion simulation **[32]**, and drug delivery modeling **[33]**. Despite widespread research, very little work has extended such transport-based modeling approaches to medical imaging, especially PET. Few studies have incorporated diffusion modeling to generate time activity curves and activity distribution maps but not synthetic images **[34–37]**. Mass transport-based solvers naturally support time-dependent dynamics, making them uniquely suited for dynamic imaging **[34]**. Additionally, the potential for patient-specific PET simulations could be explored to fill gaps in simulators for theranostic and dynamic imaging applications.

In this work, we introduce MuPET (multiphysics PET), a framework that combines diffusion–reaction modeling with a pseudo-Poisson PET noise model to generate realistic PET-like images from arbitrary geometries. We aim to validate the framework against experimental PET data and conventional lesion insertion data to demonstrate strong quantitative agreement alongside improved adaptability and computational efficiency.

## 2. Methods

### 2.1 Experimental Phantom Acquisition and Synthetic lesion Insertion

A custom imaging phantom (**Fig. 1.a**) adapted from **[26]**, was used as the physical reference system for evaluating the MuPET framework. It consists of a Jaszczak cylindrical container fitted with nine 3D-printed permeable cubes (40 mm) of acrylonitrile butadiene styrene (ABS). The cubes were designed with permeable walls to allow uniform filling and were assigned three controlled dilution levels (50%, 60%, and 70%) relative to the background solution. Six cubes contained internal spherical cavities (15 and 20 mm diameter) to emulate the lesions, whereas the remaining three were solid reference units to emulate the tissue (**Fig. 1.b**). The phantom was prefilled with a water/soap solution 4–5 hours prior to image acquisition. Fifteen minutes prior to the scan, 56.7 MBq of ^18^F-FDG radiotracer was injected into the estimated 6.2 L volume. This ensured a well-defined initial condition for both the physical measurements and the computational model. All acquisitions (**Fig. 1.c**) were performed using a single bed position on a Discovery MI PET/CT platform (*GE Healthcare, Chicago, IL, USA*) across three consecutive frames of 300 seconds each with a single CTAC (CT attenuation correction). The activity at the time of the scan was decay-corrected relative to the injected activity. PET data were reconstructed into a 384 × 384 × 89 volume with a voxel spacing of [1.042, 1.042, 2.8 mm] via Q.Clear FX BPL (*Bayesian penalized likelihood reconstruction with a regularization parameter, TOF*, β: 550). The detailed phantom preparation protocol and acquisition parameters can are followed as per guidelines by a previous study **[29]**.

**Fig. 1.**
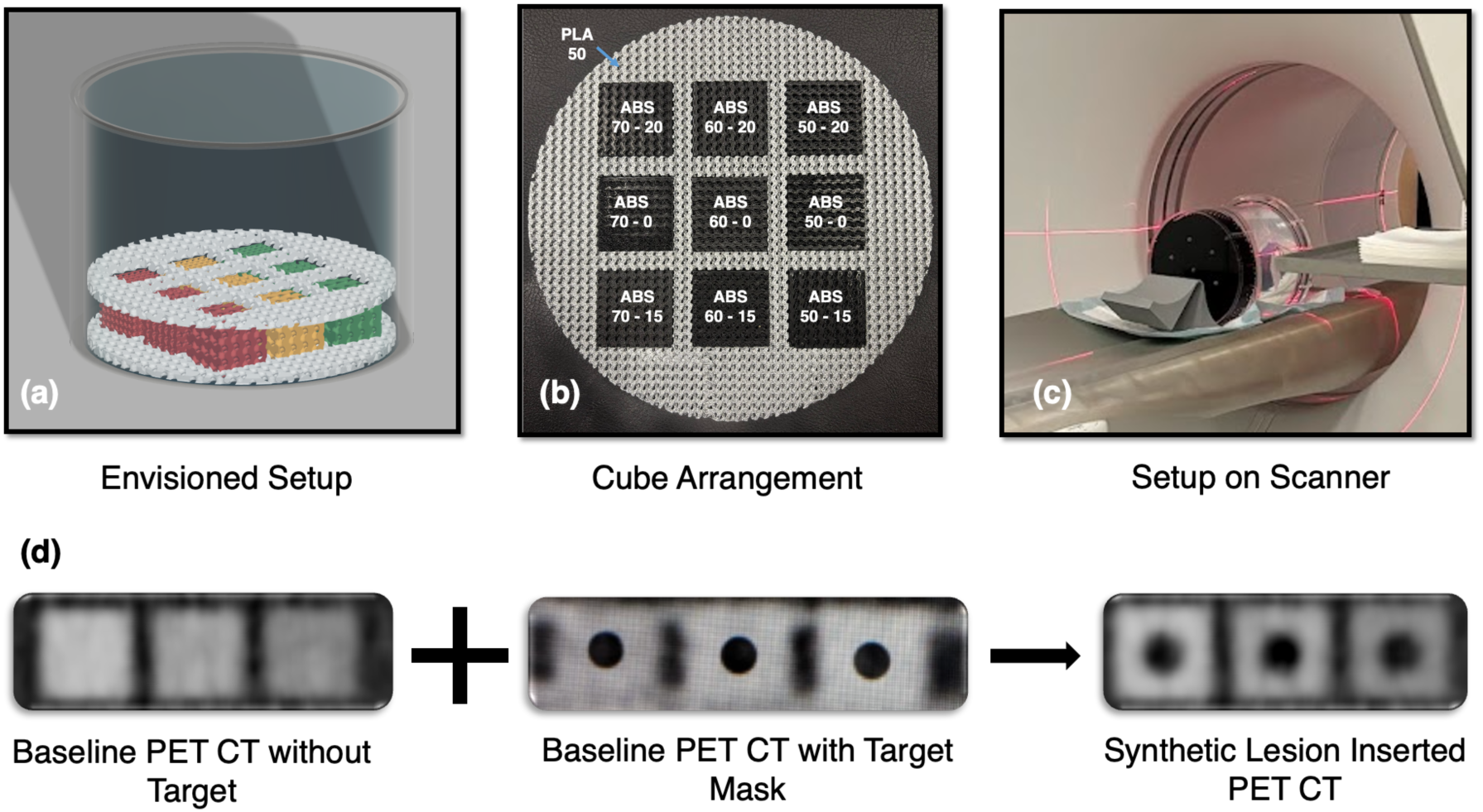
(a) 3D rendering of the adapted phantom design; (b) Arrangement of cubes – (Notation: percentage of ABS – target size); (c) Bed leveling on a PET/CT scanner; (d) Summarized workflow of synthetic lesion insertion

Synthetic lesion insertion was subsequently performed on the basis of a workflow proposed by **[14]** via the Duetto Toolbox (*GE Healthcare, Chicago, IL, USA*). This step was included to provide a benchmark against a conventional lesion-insertion workflow to enable a direct comparison with MuPET’s physically modeled synthetic data. Spherical binary masks of 15 and 20 mm were created and positioned on the reference cubes (no target) in the acquired PET images (**Fig. 1d**). Inserted synthetic lesions (ISLs) were created by specifying the target-to-background activity ratio with respect to the reference cube measurements to the generated masks, which were converted into simulated sinograms. This was added to the raw sinogram, and the final lesion-inserted images were generated through a reconstruction process and parameters matching the acquisitions.

### 2.2 MuPET Framework

The MuPET (multiphysics PET) framework introduced here presents a computationally efficient pipeline for simulating PET image data. This approach combines COMSOL Multiphysics^®^ (*COMSOL Inc., Stockholm*) based tracer distribution modeling with a synthetic noise modeling postprocessing workflow.

#### 2.2.1 Numerical Modeling

Conservative mass transport was modeled via a diffusion-reaction formulation (**Eq. 1**), assuming concentration-driven passive movement coupled with first-order radioactive decay. The simulation geometry for the numerical simulation was constructed based on experimental dimensions and measurements adjusted to measure the fillable volume, as represented in **Fig. 2a**.

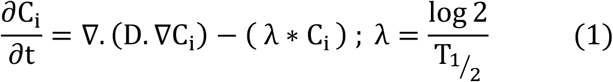

where C_i_ is the concentration in mol/m^3^, D is the diffusion coefficient (m^2^/s), λ is the decay factor (1/s), and T_1/2_ is the half-life (s).

**Fig. 2.**
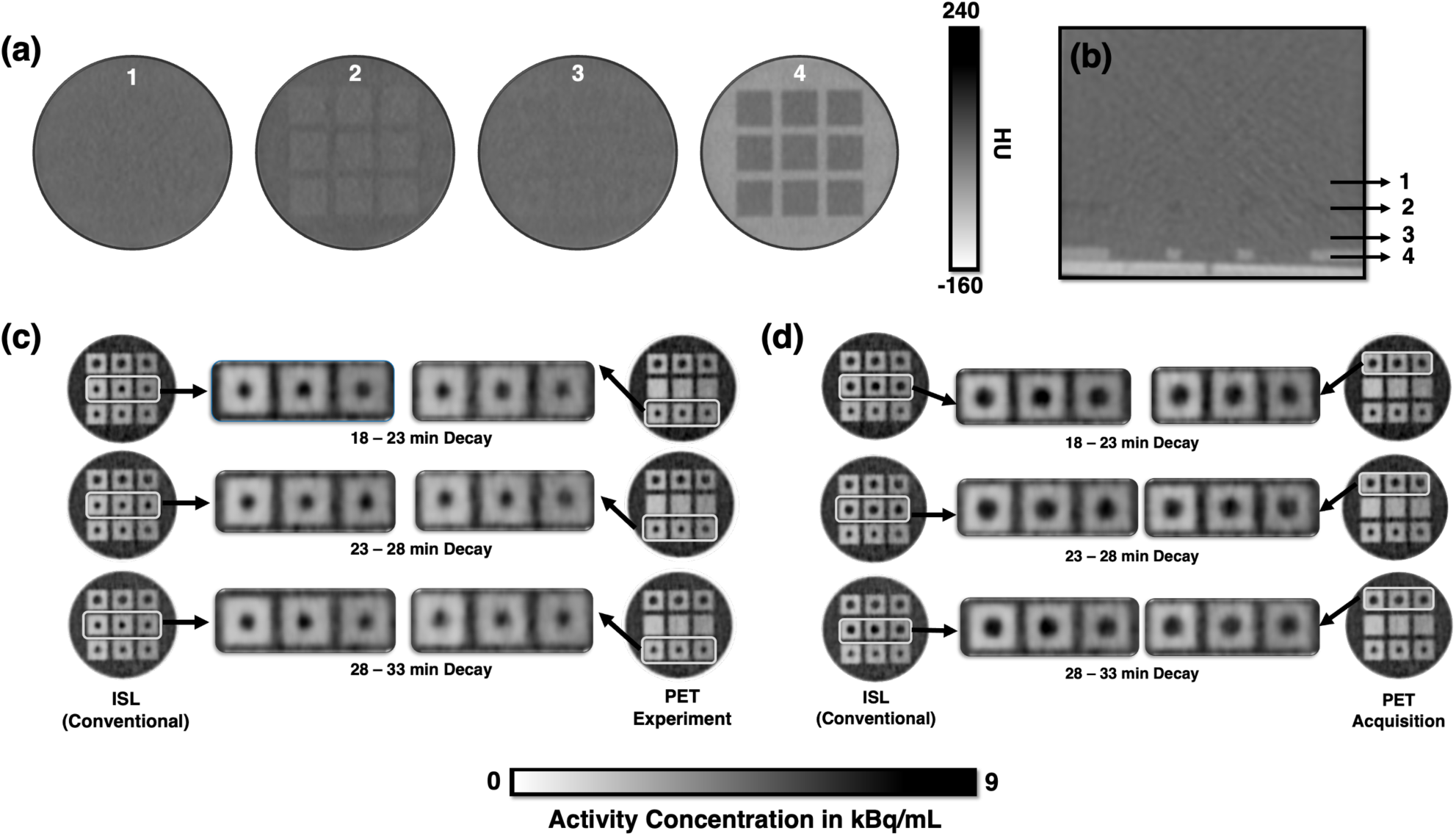
**(a)** Cross-sectional CT AC slices at 4 different locations (1. Pure water, 2. ABS cubes with ABS support, 3. Cubes with water; 4. ABS cubes with PLA support) **(b)** Corresponding location of CT slices; Central PET slice with exploded target view of ISL vs. PET acquisition for 3 frames **(c)** 15 mm, **(d)** 20 mm

The activity concentration (AC) in kBq/mL was converted to C_i_ in mol/m^3^ based on **Eq. 2**. A no-flux boundary condition was applied to the outer walls (**Eq. 3**). The initial activity concentration in the background (AC_in_) for the simulation was derived as the injected activity divided by the injection volume. Initial background concentrations AC_in_ were derived from the injected activity/volume and modulated to establish target contrast levels (i.e., 0.5 × AC_in_, 0.4 × AC_in_, or 0.3 × AC_in_, respectively).

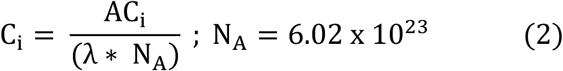

where N_A_ is Avogadro’s number in mol^-1^ and AC is the activity concentration in kBq/mL.

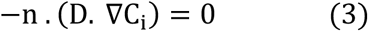

All material properties within the numerical model were assumed to be analogous to those of ^18^F-FDG and water, and in instances where specific properties were not readily available, corresponding values for glucose were adopted as a reasonable approximation. The properties used are as follows: density of the fluid (ρ): 1.00 g/mL; diffusion coefficient of FDG in water (D): 6 × 10^-10^ m^2^/s is adapted from the value of glucose **[38]**; half-life of tracer (T_1/2_): 110 minutes **[36]**. To validate the modeled geometry, volume-averaged activity (MBq) in the domains was compared with decay-corrected values from acquired image DICOM metadata. The computational domains were discretized and independence of the concentrations sampled at 726 (11 × 11 × 6) different locations, with an acceptable threshold of 5% across 3 grid sizes for numerical stability was verified. The simulations were computed for 35 minutes with a step size of 1 min via an iterative solver.

#### 2.2.2 Synthetic noise modeling

Noise modeling was performed entirely in the image domain to account for PET physics noise and scanner resolution effects and few reconstruction-induced effects.

a. The simulated activity maps AC_Simulated_ (x, y, z, t) in kBq/mL were exported as a 4D volume with spatial dimensions matching the experimental scan volumes of 383 × 383 × 89 (1.042 × 1.042 × 2.8 mm^3^). Additionally, the maximum and minimum values of activity concentration within the computational domain are exported as functions of time AC_Max_(t) and AC_Min_(t).
b. Upon import into MATLAB (*MathWorks INC, MA, USA*), postprocessing, including data cleaning and reorientation to the FOV, was performed if necessary. The volumes are also separated into individual 3D volumes.
c. 3D activity maps were converted to expected counts (CTSs) via **Eq. (4**), which includes T_scan_ (scan duration), S_scanner_ (scanner sensitivity), and V_voxel_ (voxel volume) adapted from NEMA (National Electrical Manufacturers Association) **[39]**

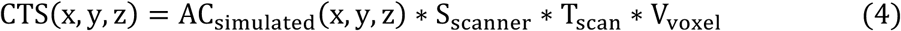

The resulting count volume CTS (x, y, z) was then subjected to a pseudo Poisson model: the counts were multiplied by an adjustable scale factor to generate CTS_(0,+._(x, y, z), after which Poisson noise was generated to introduce PET physics-based noise CTS_6_(x, y, z). Then, the noisy data were normalized back with a scale factor C_67_(x, y, z) to maintain the mean variance ratio (MVR) as K_scale_ **[20]**.

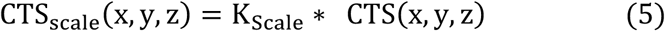

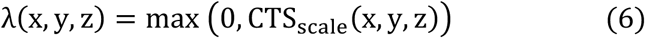

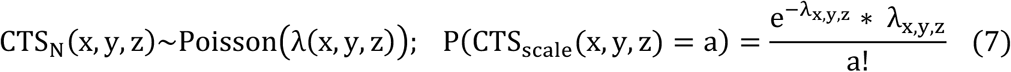

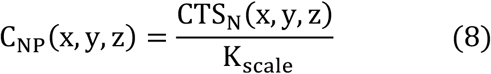

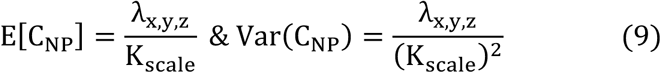

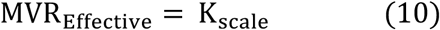

To simulate inherent resolution limitations, the AC_map_ was convolved with a spatially variant, anisotropic Gaussian kernel. The full width at half maximum (FWHM) of the kernel was derived from scanner specifications as per AAPM (American Association of Physicists in Medicine) Task Group 126 (TG-126) findings **[40]**. A detailed mathematical description and the values of the FWHM used for the Gaussian model are included in the supplementary information.

To create a scanner and acquisition-dependent dataset of the simulated noise, iterative variations of K_scale_ from 1 down to 0 were performed. For each value, a noisy volume was generated. The mean variance ratio (MVR) for each generated sample was computed via two distinct methods: 2D region of interest (ROI) extraction across six randomly chosen uniform slices and 3D volume of interest (VOI) extraction for a 75 mm uniform activity section from the center to the top of the phantom. On the basis of the extracted noise characteristics, the best fit between the calculated MVR values and their corresponding K_Scale_ values was performed.

### 2.3 Data extraction and statistical tests

The PET volume was categorized into 3 different regions: reference water, background cubes and targets for data extraction. Quantitative data extraction was carried out via the VOI (matching physical dimensions of the target) approach following the workflow of **[14,26]** on Image Interpretation Software - Volume Viewer, PETVCAR® (*GE Healthcare, Chicago, IL, USA*) for experimental and ISL data, MATALB for MuPET. The activity values in kBq/mL were extracted semiautomatically. Additional ROIs (30 mm for cubes, 20 mm for reference water) and central activity profile extractions were performed for distribution analysis. For the ROI, a total of 13 circular ROIs—9 (< 30–9 cubes) and 4 (< 20–4 background water)—were placed automatically on the central cross-sectional slice with the targets. Two quantitative metrics were calculated from the extracted VOI data, namely, the recovery coefficient (RC), which was used to quantify the recovered true activity concentrations, and the target contrast ratio (TCR), which was used to assess the contrast of lesions against the background (Eqs. 9 & 10).

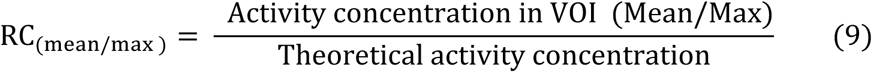

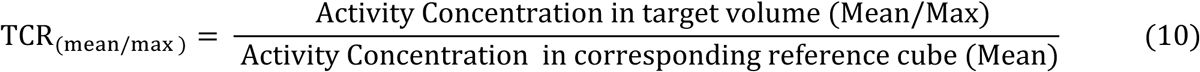

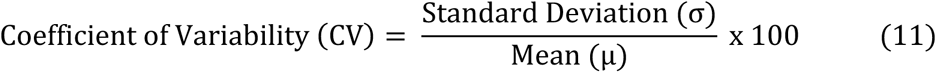

The coefficient of variability for the uniform regions was also calculated via Eq. (11) based on the extracted VOI data of acquisition and MuPET. Detailed statistical tests for the 3 quantitative metrics were not performed because of the lower target sample size of N_Sample_ = 6 for each set, and the standard percentage error and Bland‒Altman bias with limits of agreement were calculated. All the extracted activity data were initially checked for normality via the Shapiro‒Wilk test. This was followed by a two-sided Brown test and *equal* variance test on the ROI data of acquisition (Acq), and MuPET was chosen because of its reduced dependence on normality. Finally, the activity profiles for MuPET vs Acq and MuPET vs ISL were followed up by Wilcoxon rank sum test (also known as the

Mann‒Whitney test) for the assessment of central tendency-driven significant differences. All tests were performed with a significance of p < 0.05 in JASP (*JASP Team, Netherlands*).

## 3. Results

### 3.1. Phantom Acquisition and Synthetic Lesion Insertion

The observed Hounsfield units (HUs) of the CT values of the reference cubes were 4.6 ± 4.8 (TBR ∼ 2), 4.5 ± 3.5 (TBR ∼ 2.5), and 7.6 ± 4.3 (TBR ∼ 3.33), which fell within the range of water (3.1 ± 3.7) (**Fig 2.a, 2.b**). All the different regions were captured effectively via PET acquisition (PET Acq), with different contrasts being visibly distinguishable (**Fig 2.c, 2.d)**. Additionally, the inserted synthetic lesions (ISLs) represented the actual targets well enough, and a visual comparison based on the target diameter was performed (**Fig 2.c, 2.d).**

### 3.2. Mass Diffusion and Decay Reaction Simulation

The computational phantom, as depicted in **Fig. 3.a**, was estimated as a 6.23 L volume. The effective activity (MBq) calculated from the volume-averaged simulated activity concentration was compared against the analytically decay-corrected experimental values (DICOM metadata) (**Fig. 3.b**). The percentage differences observed for the activity were low, ranging only from 0.43% to 1.16%, confirming high model fidelity. Numerical stability was validated through a grid convergence study **(Fig. 3.c).** Effective convergence was achieved between 0.6 and 1 million tetrahedral elements, and a final mesh of 1 million tetrahedral elements was selected for all subsequent analyses. This specific configuration resulted in a computational time of ∼ 10 minutes for a total simulated decay of 35 minutes (with a time step of 1 minute). The central cross-sections of the activity concentrations for the 3 corresponding time steps are shown in **Fig.** 3d, with observed maximum–minimum values of 8.26–2.16, 8.10–2.13, and 7.84–2.09 kBq/mL (averaged across 3 frames).

**Fig. 3.**
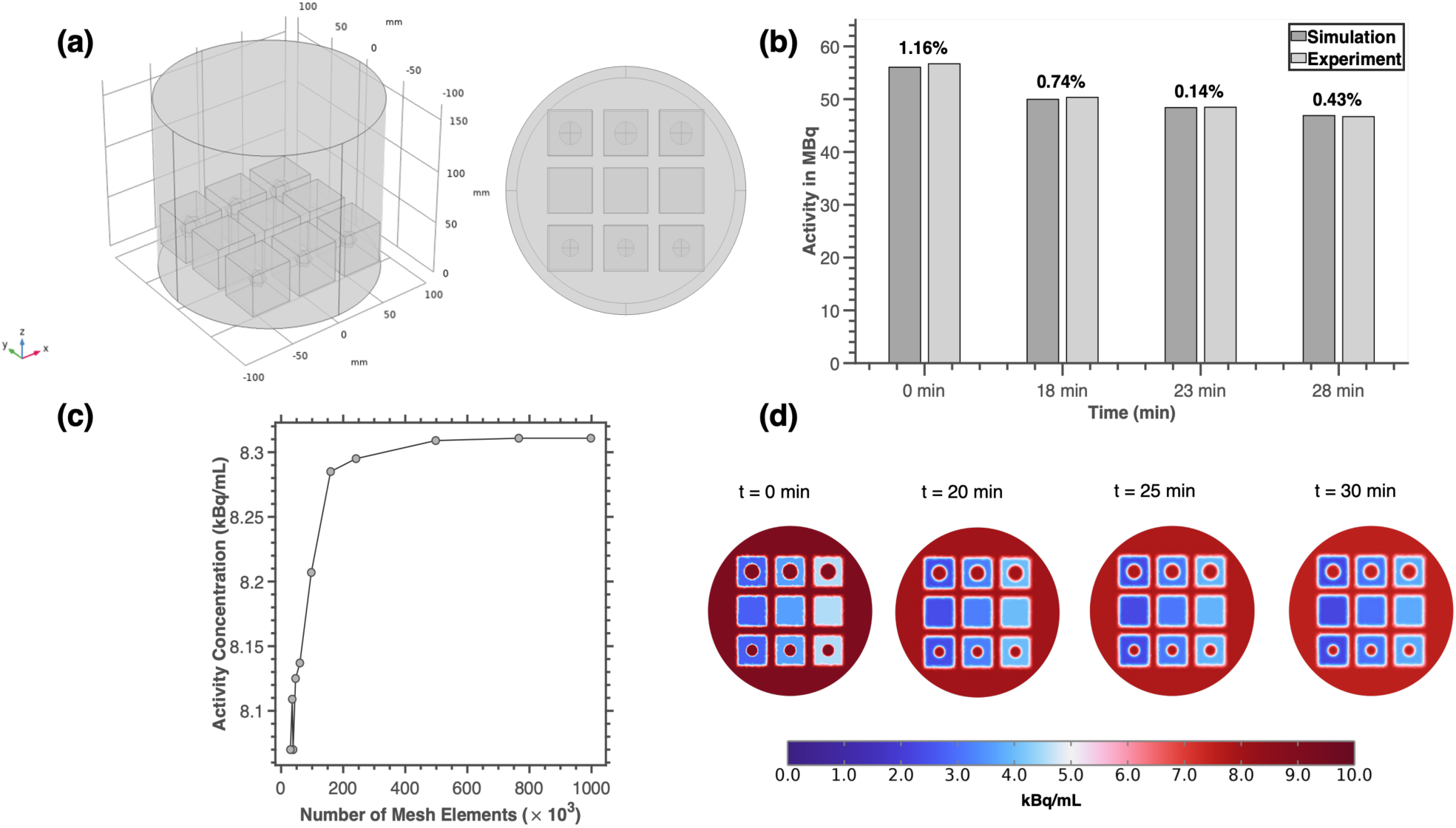
**(a)** 3D rendering of the computational domain and axial (top view) geometric setup displaying targets and cubes following the experimental layout. **(b)** Comparison of the effective activity (in MBq) of the simulation with analytically calculated decay-corrected activity from DICOM metadata. **(c)** Grid convergence for the simulation. **(d)** Central cross-sectional slice for the simulated activity distribution at different time stamps prior to noise addition.

### 3.3. Noise characterization and MuPET modeling

For the PET noise model, the MVR_acquisition_, which was determined from scan data on the basis of a central VOI measuring <λ 18 cm × 7.5 cm, yielded MVR = 85.8. Additionally, MVR values extracted from ten random uniform slices utilizing an 18 cm ROI **[19]** resulted in a consistent range of values between 75 and 95. The pseudo-Poisson noise model for simulation was calibrated by varying the scale factor K_scale_ between 0.005 and 1. **Fig. 4a,b** shows samples of the simulated noise, illustrating how this parameter influences the noise distribution. This process established a linear relationship between the resulting MVR_Noise_ and K_scale_ (**Fig. 4.a**). Notably, this fit is intrinsically specific to the scanner properties and Gaussian used in the scanner model. Using the linear fit, a global K _scale_ of **0.0415** was selected, yielding a resultant MVR_sim_ = 85 thus calibrated to acquisition value. This optimized noise model resulted in coefficient of variation (CV) values for MuPET (3.9-4.04%) that closely matched the acquired PET data (3.75-3.91%).

**Fig. 4.**
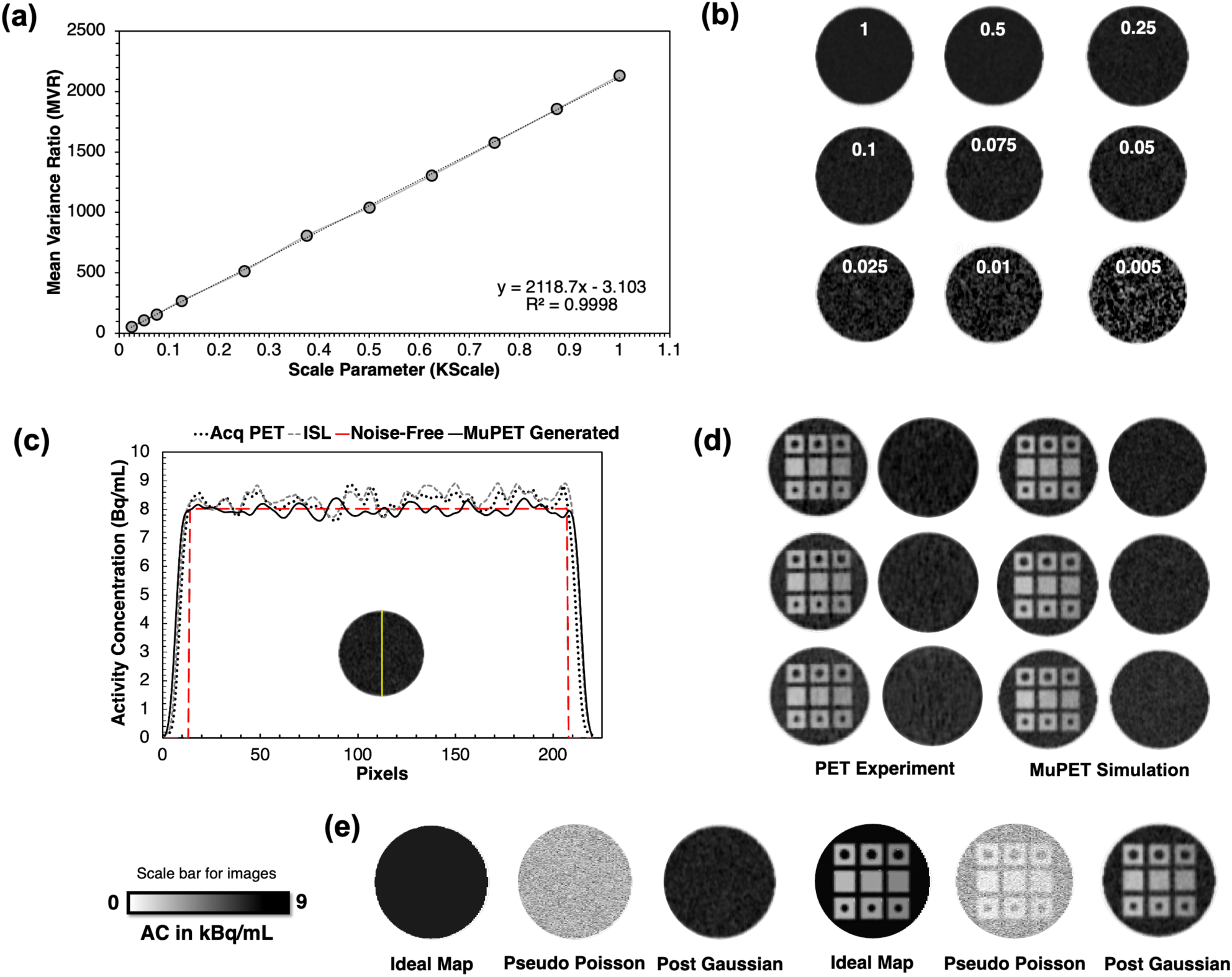
**(a)** Linear fit for K_Scale_ as a function of MVR for the simulation map. **(b)** Cross-sectional slice of generated noise for different _KSc._ **(c)** Activity profile extracted experimental data, simulated map and corresponding generated noise for frame 1. **(d)** Background and target images of the 3 frames - Acquisition PET (COV_BKG_ – 3.75, 3.83, 3.91%) vs MuPET simulation for K_Scale_ ∼ 0.03 (COV_BKG_ – 3.63, 3.64, 3.75%). **(e)** Overview of noise generation workflow for background and target slices.

### 3.4. Quantitative comparison of PET profiles and metrics

The extracted ROI data distributions were compared via kernel density estimation (KDE) (**Fig. 5**). The activity profiles extracted along the central vertical axis are presented in **Fig. 6a**. The target profiles exhibited a Gaussian-like peak, characteristic of partial volume effects (PVEs), confirming the integrated resolution blurring effect. In addition to visual assessment, quantitative metrics provide deeper insight into the model’s performance. The recovery coefficients and target contrast ratios averaged over 3 frames are presented in **Fig. 6.b, 6.c & 6.d**. Bland‒Altman bias analysis was used to quantify the differences between methodologies (**Table 1**). Compared with acquired PET, MuPET tended to slightly underestimate the maximum values (RC_Max_, TCR_Max_), whereas the ISL method generally overestimated these metrics. All the mean values were within a comparable range of bias of 10% of acq.

**Fig. 5.**
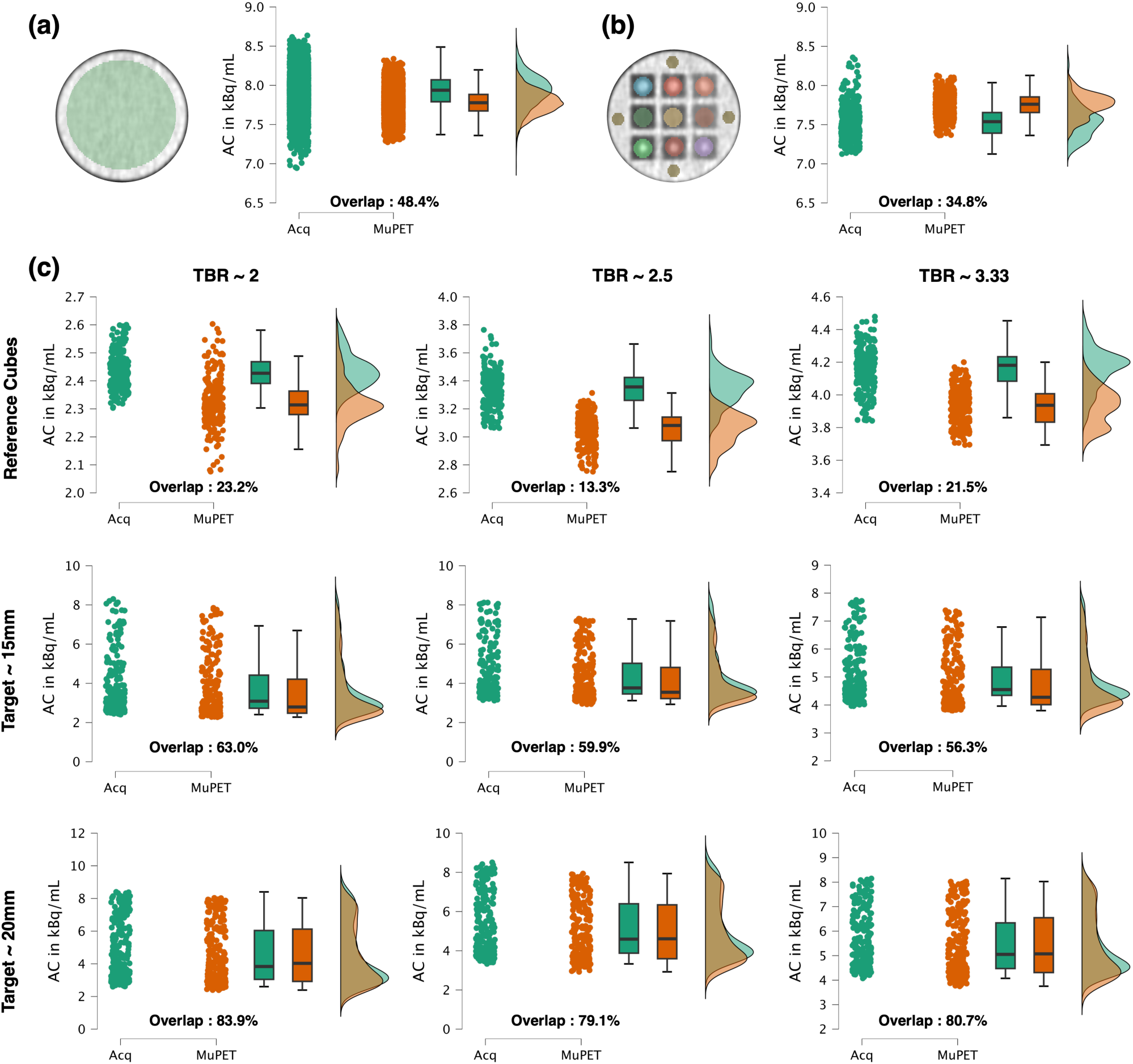
**(a)** Extracted activity distribution for the 18 cm ROI on the uniform slice for MuPET vs Acq; **(b)** ROI placement (30 mm for cubes, 20 mm for water) and corresponding data distribution comparison for water ROIs; **(c)** Corresponding comparison of raincloud data distribution plots for cubes - reference cubes, 15- and 20-mm targets (rows); TCR ∼ 2.00, 2.50, 3.33 (columns).

**Fig. 6.**
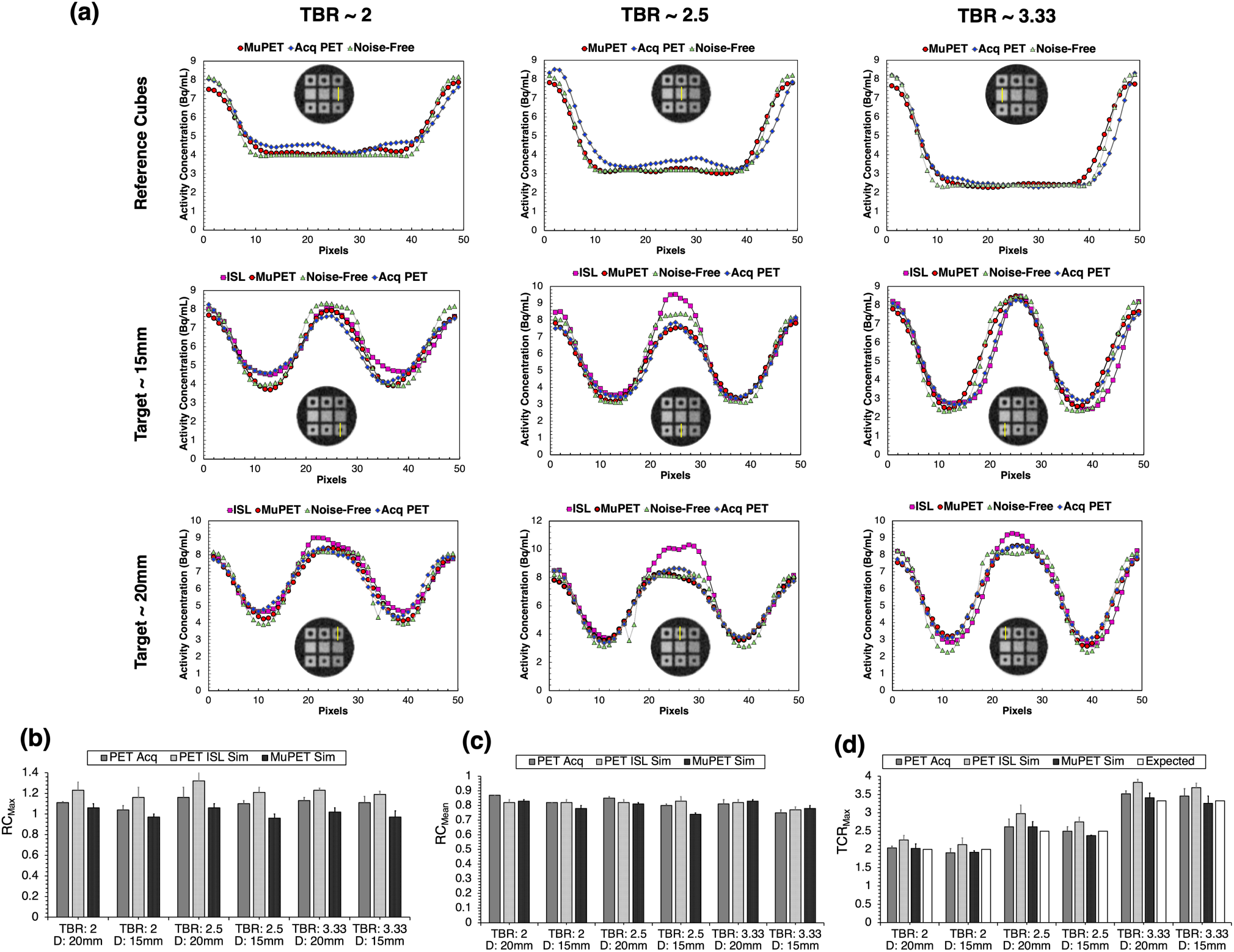
**(a)** Vertical activity profile of cubes on a central cross-sectional slice – reference cubes and 15-and 20-mm targets (rows); TCRs ∼ 2.00, 2.50, and 3.33 (columns). Averaged quantitative metric comparison based on VOI data for **(b)** RC_Mean_, **(c)** RC_Max_, and **(d)** TCR_Max_ (the corresponding data can be found in the supplementary information).

**Table. 1.** Bland‒Altman mean bias percentage with limits of acceptance (mean ± 1.96 * SD) for quantitative metrics.

| Quantitative<br>Metric | Acq vs ISL | Acq vs MuPET | ISL vs MuPET |
| --- | --- | --- | --- |
| $RC_{Max}$ | $-9.84 \pm 4.02$ | $9.64 \pm 6.98$ | $10.27 \pm 4.04$ |
| $RC_{Mean}$ | $0.33 \pm 7.29$ | $2.66 \pm 9.72$ | $2.32 \pm 9.99$ |
| $TCR_{Max}$ | $-9.73 \pm 4.29$ | $2.34 \pm 5.37$ | $12.06 \pm 2.94$ |

The Brown-Forsythe test indicated no statistical evidence of variance (**Table 2**) for most conditions, except for the 50% reference cube and the uniform slice.

**Table. 2.** Summary of descriptive data & nonparametric statistical tests.

|  |  | TCR ~ 3.33:1 |  |  | TCR ~ 2.50:1 |  |  | TCR ~ 2.00:1 |  |  | Background |
| --- | --- | --- | --- | --- | --- | --- | --- | --- | --- | --- | --- |
|  |  | R | 15 mm | 20 mm | R | 15 mm | 20 mm | R | 15 mm | 20 mm |  |
| ROI Data | Mean in kBq/mL (Coefficient of Variability %) |  |  |  |  |  |  |  |  |  |  |
|  | Acq | 2.44 (3%) | 3.79 (40%) | 4.62 (39%) | 3.35 (4%) | 4.42 (30%) | 5.17 (29%) | 4.16 (3%) | 4.98 (19%) | 5.49 (22%) | 7.92 (3%) |
|  | MuPET | 2.32 (4%) | 3.58 (42%) | 4.56 (39%) | 3.06 (4%) | 4.13 (30%) | 4.98 (31%) | 3.93 (3%) | 4.74 (21%) | 5.44 (24%) | 7.78 (2%) |
|  | Brown Forsythe P – value: N<sub>Sample</sub> ~ 17000 (For background), 990 (For rest) |  |  |  |  |  |  |  |  |  |  |
|  | - | 0.01 | 0.75 | 0.85 | 0.45 | 0.69 | 0.63 | 0.63 | 0.49 | 0.08 | 0.001 |
| Wilcoxon Rank Sum P – value: N<sub>Sample</sub> ~ 442 (For background), 98 (For rest) |  |  |  |  |  |  |  |  |  |  |  |
| Activity Profile (MuPET vs Acq) |  | 0.91 | 0.79 | 0.80 | 0.001 | 0.97 | 0.72 | 0.008 | 0.69 | 0.54 | 0.05 |
| Activity Profile (ISL vs Acq) |  | - | 0.45 | 0.99 | - | 0.23 | 0.19 | - | 0.28 | 0.27 | 0.00135 |

## 4. Discussion

The MuPET framework, as introduced in this work, successfully integrates diffusion-reaction modeling with an empirical noise model and resolution blurring to generate realistic synthetic PET data. In this work, we validated our method exclusively via controlled phantom experiments. This choice was intentional, as phantoms provide a fully controllable and well-defined ground truth against which the underlying physics model and noise pipeline can be rigorously assessed. Model fidelity despite several approximations is validated by strong quantitative agreement between the simulated effective activity and the decay-corrected experimental values, with a variability of < 5%.

Our mass diffusion model effectively represents the tracer distribution. This is crucially validated by the qualitative and quantitative agreement between the simulated effective activity (**Fig. 3.b**) and the noise-free target activity profiles (**Fig. 3.d**), which follow experimental patterns.

A key computational simplification is the assumption of bulk concentrations instead of modeling microlevel tracer uptake. This approximation is validated because the phantom’s microlevel structural variations (pores typically < 4 mm) are smaller than the scanner’s resolution. Consequently, the PET image captures only the average macroscopic tracer behavior, allowing the model to focus on bulk simulation while reducing the computational intensity by avoiding high refinement zones.

We assume that the diffusion properties of the ABS cubes are identical to those of water + FDG. This choice is radiologically justified by the HU values **[29]** observed for the reference cubes in the CT acquisition (Section 3.1), which fall within the range of water. However, the true diffusivity in the cubes is much lower than that in the bulk water because of microscale geometrical variations. Although not observed in this study, the use of water properties in simulations risks overestimating diffusion over extended periods. In future, longer diffusion study is needed to assess scattering fully. A robust solution in this regard could be is to build an empirical dataset following homogenization principles: microscopic changes in permeability and diffusivity are mapped to a macroscopic value as a function of porosity. With respect to the pseudo scaled Poisson noise model, the primary goal was to mimic quantitative stochastic noise rather than voxel-based textural changes. In this regard, the models global MVRsim 85 resulted in CoV_sim_ values (3.9-4.04%) closely matching the acquired PET values (3.75-3.91%). However, some discrepancies can also be observed in the ROI data (**Table 2 & Fig. 5**). Across all evaluated target sizes, MuPET showed strong alignment with both acquisition and ISL for TCR_max_ and RC_mean_. This is also supported by the close activity profiles (**Fig. 6.b**) to those of the experiment and ISL. The resulting p values also indicate the absence of statistical evidence, although with some outliers. To assess partial volume effects (PVEs) accurately, a detailed analysis using common segmentation criteria, rather than 2D profiles, would be ideal for future work.

Notably, the conventional synthetic lesion insertion method consistently overestimates the values, especially RC_max_ and TCR_max_. This is because applying the contrast ratio to a base experimental scan, which inherently carries PET noise multiplies the bias. In contrast, the results of the MuPET noise model were slightly underestimated compared with those of the experiment (**Table 2**), which can be attributed to two main factors: the simulation being instantaneous while experimental scans are acquired over time and the global application of the noise scaling parameter (K_scale_) despite noise naturally increasing as activity concentration decreases. The former can be addressed by selecting the last timestamp or an average across time stamps corresponding to the scan duration instead of an intermediate time point. The latter requires that further iterations incorporate the dependence of K_scale_ on voxel intensity. Additionally, while we followed the noise characterization approach of **[19]**, the MVR can be sensitive to the chosen ROI or VOI; a more robust analysis, such as noise power spectrum (NPS) analysis **[17]** or automatic slice-by-slice noise mapping **[41],** could better determine the noise characteristics. Another interesting phenomenon observed was that the overlap between the extracted ROI activity values was the worst for the reference cubes, and the target cubes provided the best representation. This accurate depiction of lesions is particularly promising considering that our noise model was generated based on uniform, nontarget slices. The higher COV for the target cubes (**Table 2**) is because the ROI (30 mm) includes the target and certain regions of background to obtain consistent measurements across methods for comparison. In this framework, the mean values are generally affected by the simulation, and the variance is affected by the noise model. The underestimation observed in low-activity reference cubes is due to the lack of voxel intensity-based effects, i.e., the BPL algorithm used for acquisitions actively suppresses noise in these regions, which is not captured by the simple noise model. Hence, further noise model datasets are needed for various reconstruction settings.

In this work, the core objective was to introduce a fast, scalable computational pipeline centered on modeling tracer distributions. The MuPET pipeline achieves this with a total computation time of less than 15 minutes (activity map simulation to PET data generation) without a baseline PET scan while the ISL took around 12h with a baseline-scan. MuPET is not intended to be a replacement for high-fidelity clinical simulators or lesion insertion methods that can model the entire scan process but rather a new pragmatic approach for quick ground truth datasets. An interesting addition for future works would be a pipeline with an approximated CT attenuation map built from 3D models **[42]**. This enables the potential to include more reconstruction algorithms, thus reducing empirically derived relationships and opening avenues for comprehensive image quality studies. The adaptability of analogous MuPET frameworks for other imaging modalities can also be explored. The core strength lies in leveraging mass transport solvers like those used in cardiovascular modeling, which naturally support time-dependent dynamics. This opens the possibility to extend future works into complex, patient-specific simulations and dynamic imaging ground truths with better noise models.

## Conclusion

The MuPET framework successfully integrates multiphysics transport modeling with image-domain noise simulation to produce realistic synthetic PET data. Validated against physical phantom experiments, the method accurately predicts tracer distributions with high fidelity while replicating clinical noise characteristics. By utilizing mass transport solvers, MuPET establishes a practical foundation for future extensions into dynamic PET Image simulation.

## Data Availability

All data produced in the present study are available upon reasonable request to the authors

## Abbreviations used in the article

**Abbreviations used in the article:**
|  |  |
| --- | --- |
| **AAPM** | American Association of Physicists in Medicine |
| **ABS** | Acrylonitrile Butadiene Styrene |
| **AC** | Activity Concentration in kBq/ml |
| **Acq** | Acquisition |
| **ANOVA** | Analysis of Variance |
| **BPL** | Bayesian penalized-likelihood reconstruction |
| **C<sub>i</sub>** | Concentration along direction i in mol/m<sup>3</sup> |
| **COV** | Coefficient of Variability |
| **CTAC** | Computed Tomography Attenuation Correction |
| **CTS** | Counts |
| **D** | Diffusion Coefficient |
| **DICOM** | Digital Imaging and Communications in Medicine |
| **DMI** | Discovery MI |
| **EM** | Expectation-maximization |
| **FAST PET** | Fast analytical simulator of tracer PET |
| **FDM** | Fused Deposition Modeling |
| **FDG** | Fluorodeoxyglucose |
| **FOV** | Field of View |
| **FWHM** | Full Width at Half Maximum |
| **HU** | Hounsfield Units |
| **ISL** | Inserted Synthetic Lesion |
| **KDE** | Kernel Density Estimation |
| **K<sub>scale</sub>** | Noise Scale Parameter |
| **MuPET** | Multiphysics PET |
| **MVR** | Mean to Variance Ratio |
| **n** | Normal Vector |
| **N<sub>A</sub>** | Avogadro Number |
| **NEMA** | National Electrical Manufacturing Association |
| **PET/CT** | Positron Emission Tomography/Computed Tomography |
| **PETSTEP** | PET Simulator of Tracers via Emission Projection |
| **PSF** | Point spread function |
| **PVE** | Partial volume effects |
| **QC** | Quality Control |
| **RC** | Recovery Coefficient |
| **ROI** | Region of Interest |
| **SMART PET** | SiMulAtion and ReconsTruction PET |
| **S<sub>scanner</sub>** | Scanner sensitivity in cps/kBq |
| **T<sub>1/2</sub>** | Half Life of Radiotracer in min |
| **TCR** | Target Contrast Ratio |
| **TG 126** | Task Group 126 Report |
| **TOF** | Time of Flight |
| **T<sub>scan</sub>** | Scan Duration |
| **VOI** | Volume of Interest |
| **V<sub>voxel</sub>** | Voxel Volume in mL |
| **λ** | Decay factor in s<sup>-1</sup> |
| $\mu$ | Mean |
| $\sigma$ | Standard Deviation |
| $\rho$ | Density of radiotracer in g/mL |
| $\phi$ | Diameter |

